# Development and Evaluation of an Android-based Platform for Early MCI Detection in an Elderly Population

**DOI:** 10.1101/2020.03.16.20037028

**Authors:** Mahsa Roozrokh Arshadi Montazer, Roohollah Zahediannasb, Roxana Sharifian, Mahshid Tahamtan, Mahdi Nasiri, Mohammad Nami

## Abstract

**Background:** Mild cognitive impairment (MCI) is an intermediate stage of cognitive decline fitting in-between normal cognition and dementia. With the growing aging population, this study aimed to develop and psychometrically validate an android-based application for early MCI detection in elderly subjects.

**Method:** This study was conducted in two phases, including 1-Initial design and prototyping of the application named M-Check, 2-psychometric evaluation. After the design and development of the M-Check app, it was evaluated by experts and elderly subjects. Face validity was determined by two checklists provided to the expert panel and the elderly subjects. Convergent validity of the M-Check app was assessed using the Montreal Cognitive Assessment (MoCA) battery through Pearson correlation. Test-retest and internal consistency and reliability were evaluated using Intra-Class Correlation (ICC) and Kuder-Richardson coefficients, respectively. In addition, the usability was assessed by a System Usability Scale (SUS) questionnaire. SPSS 16.0 was employed to analyze the data.

**Result:** The app’s usability assessment by elderlies and experts scored 77.11 and 82.5, respectively. Also, the correlation showed that the M-Check app was negatively correlated with the MoCA test (r = -0.71, p <0.005), and the ICC was more than 0.7. Moreover, the Richardson’s Coder coefficient was 0.82, corresponding to an acceptable reliability.

**Conclusion:** In this study, we validated the M-Check app for the detection of MCI based on the growing need for cognitive assessment tools that can identify early decline. Such screeners are expected to take much shorter time than typical neuropsychological batteries do. Additional work are yet to be underway to ensure that M-Check is ready to launch and used without the presence of a trained person.

## 1. Introduction

Dementia is a syndrome with deterioration in memory, thinking, behavior, and the ability to execute daily activities mostly concerned in older people yet not a normal part of aging(1). The rate of dementia is predicted to increase affecting 46.8 to 131.5 million people between 2015 and 2050 (2–4). One predominant cause for dementia is Alzheimer’s disease (A for which no specific treatment is yet available (2,3).

Alzheimer’s disease is a type of degenerative brain disorder which tend to progress over time. The process is estimated to occur 20 years or more before symptoms arise (5). The prevalence rates for AD also increase with age, especially after the age 65. Indeed, the prevalence of dementia, especially AD, is subject to 15-fold increase between the age 60-85 years (6). According to some reports, the total cost burden for medical care of individuals with Alzheimer’s or other dementias in the United States has exceeded $290 billion in 2019(7). Hence, early detection of Alzheimer’s and other dementias provides many advantages for AD sufferers and their families including medical, social, emotional, and health care planning benefits(7). As such, early detection of Alzheimer’s dementia in prodromal stages such as mild cognitive impairment (MCI) would potentially lead to proper care planning and provision with subsequent decrease in disease progression rate.

MCI is an intermediate stage of dementia (8) fitting in between normal cognition and dementia (9). The prevalence of MCI in adults older than 65 years is between 10% and 20 %(10). Impaired cognitive skills can affect social, functional, and occupational activities (11) leading to consequent problems for families and patients (12). Cognitive change can be combined with functional impairments, further deteriorates the quality of life (13,14). An increase in life expectancy leads to a growing aged population. As such, it appears that the assessment of cognitive functioning over time is crucial for the timely detection of cognitive impairment. The traditional neuropsychological battery is burdensome, time-consuming, requiring expert interpretation (15). Raising public awareness towards differentiating MCI from healthy aging would help timely evaluations.

In this vein, some online and computer-based screening tools have been established for dementia and related condition. While some of such platforms have been developed and validated from scratch, others have emerged from the traditional examination tools (16). Smartphones are expected be employed as one of the main platforms for the extension of clinical care/research in our digital world (17). The utilization of mobile health in the elderly population can be useful in enhancing care, health-management, self-efficacy, health-related behaviors such as sleep, diet, physical activity, and mental health, as well as adherence to medication (18). The use of mobile technologies in the assessment of cognitive and psychological domains in the elderly can also be validated for use as reliable tools (19).

With respect to self-evaluation through mobile health, smart phones seem to provide better, faster, and cheaper services for the elderly (20).

On the other hand, due to the increasing aging population in the world, developing cognitive health applications can potentially provide general information and identify people with a likelihood of MCI. The present study has been attempted to take primary steps in cognitively screening the elderly people for MCI. The aim of this study was to develop and validate an android-based application for early MCI detection in elderly subjects.

## 2. Methods

This study was conducted in two phases including: 1-Initial design and prototyping the application named M-Check, 2-psychometric evaluation (see Figure1).

**Figure 1:**
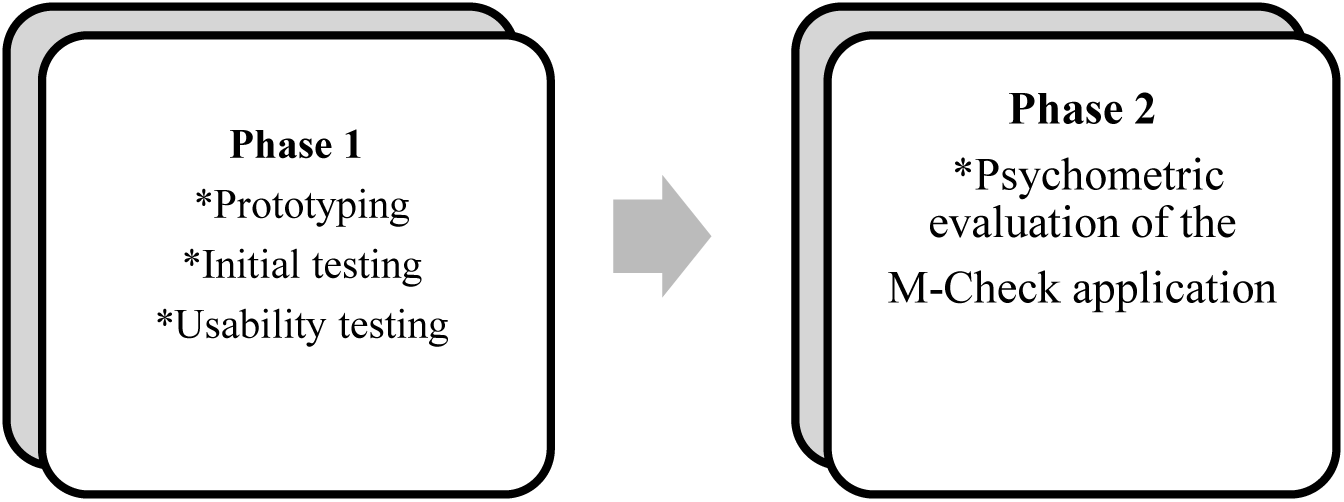
Diagram of the research phases

### 2.1. Phase 1: Initial design and prototyping

The application modules including general information, demographic data, privacy statement, and other features such as layout and appearance were determined following consultation with a panel of experts (comprising neuroscience, cognitive science and medical informatics experts).

Taking the expert panel advice into consideration a prototype of the memory check (M-Check) application was developed. After several team meetings the prototype was discussed and optimized. A computer programmer was asked to codify the application (app) in Hypertext Preprocessor (or simply *PHP*) language using the ionic framework.

MySQL database was also used. The app was based on both Android Studio 2.1.2 and website environments. The program compilation was done using the Android version 5. After initial testing, the app was debugged until the desired features were obtained.

#### 2.1.1. Application modules

The M-Check app consisted of six main modules including general information, performing Cognitive Impairment Test, status chart, contact us, link to Brain Health Centers in Iran, and the app evaluation section (Figure 2).

**Figure 2:**
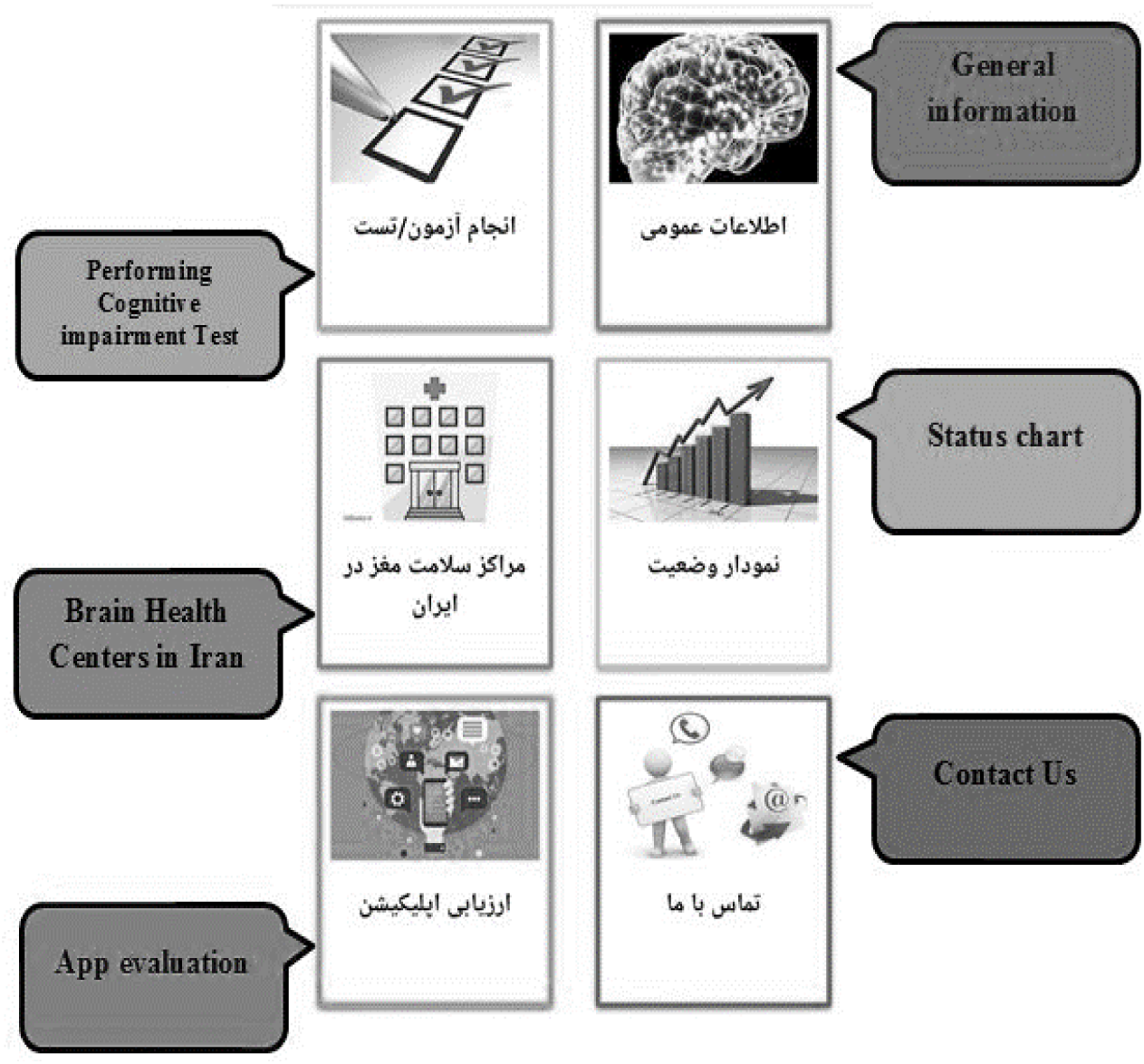
Application modules

#### 2.1.2. General information

Researchers extract general information describing the dementia process, benign forgetfulness, subjective cognitive impairment (SCI), MCI and its signs and symptoms, diagnosis and treatment of MCI, risk factors associated with MCI, Alzheimer’s dementia and the proper time to seek medical advice. The above information was recompiled from relevant peer-reviewed published papers (2000-2020) and translated into Persian.

#### 2.1.3. Performing Cognitive impairment Test

This section comprised demographic information such as birth date, handedness, cigarette and alcohol use, physical activity, history of illnesses and drug use, family history of AD and MCI assessment. The assessment module for MCI was based on the Alzheimer’s Dementia Questionnaire (21) which was validated with the reliability of its paper-based format assessed in the previous study. This app is designed for both self-assessment and other-evaluation purposes so that people can assess themselves if they are able to do the test; otherwise, each family member could complete the test for the individual. At the end of the test, based on the total score, the appropriate outcome message were presented as follow:

- *Score 0 to 4:* Your test result is within the normal range. You can improve your brain function and cognitive capacity through recommended cognitive empowerment trainings.
- *Score 5 to 14:* The result of your test may indicate symptoms of mild cognitive impairment. It is then advisable you refer to brain health centers for further evaluation.
- *Score 15 to 28:* The result of your test may indicate more severe cognitive predicament. It is then advisable you refer to the brain health centers at earliest.

#### 2.1.4. Status chart

Individuals could see the trend of change in their cognitive status according to sequential self-assessment scores.

#### 2.1.5. Brain Health Centers in Iran

This section links the audience to brain health centers in Iran where indicated. That way, individuals can refer to such centers in their city or province where applicable.

#### 2.1.6. Contact Us

In this section, communication channels with the developers were provided (e.g., telephone, email, website, and physical address).

#### 2.1.7. App evaluation

Some questions were asked to assume the users’ experience on usability for consideration in future updates.

#### 2.1.8. Usability testing

Usability evaluation was performed by the system usability scales (SUS) by six experts (a psychologist, two neuroscientists, a health information management expert, medical informatics, and an information management system expert) and 13 elderlies.

### 2.2. Phase2: Psychometric evaluation of M-Check application

In this phase, face and concurrent validity, test-retest, and internal reliability of the application was assessed.

#### 2.2.1. Face validity

The research team designed a checklist based on a visual analog color scale comprising 14 questions (0-10 where a score of 0 indicated lack of the stated feature and score of 10 indicates the highest rating). Six experts and ten elderlies participated in this evaluation.

#### 2.2.2. Concurrent validity

The MoCA battery was used to assess convergent validity of the M-Check (n=76).

#### 2.2.3. Reliability

Thirty elderly subjects participated upon the evaluation of test-retest reliability of the M-Check app. First, subjects performed the test via the app, then at a two-week interval, 16 of them were retested by the paper-based version, and 14 retested by app again. Furthermore, to assess internal consistency reliability, 70 older adults took part.

#### 2.2.4. Study context and venue

The present study was conducted in adult day-care centers, cultural centers and elderly community parks located in Shiraz, Iran. Among 95 elderly individuals who took part, 25 of them were excluded for not meeting our inclusion criteria. As such, the final sample included 70 subjects. Participants were included if they had aged 60 years or older, knew how to handle smartphone with Android operation system (version 2.3.3 or higher), were able to speak and communicate, and had no significant cognitive impairment or diagnosed with dementia. Also, participants were excluded from the study if they did not tend to participate in the survey, used neuropsychiatric medications, or had a history of neurological disease, severe mental illness, and a history of traumatic brain injury.

### 2.3. Instruments

#### 2.3.1. Montreal cognitive assessment

The Montreal Cognitive Assessment (MoCA) is a brief 30-question test which helps in assessing people for dementia. The MoCA includes eight main domains i.e. Attention and Concentration, Executive Functions, Memory, Language, Visuospatial Skills, Conceptual Thinking, Calculations, and Orientation. The total score ranges from 0 to 30, with a score of 26 and higher generally considered as normal (22).

#### 2.3.2. Alzheimer questionnaire

AQ is 21 yes/no question tool with five main domains i.e. Memory, Orientation, Functional Ability, Visuospatial Ability, and Language. AQ score ranges from 0 to 27 which higher score indicating greater impairment (21). In this study, the Persian version of AQ was used in the M-Check application.

#### 2.3.3. System usability scale (SUS)

SUS is one of the usability assessment tools developed by John Brooke. This tool consists of ten questions rated on a 5-point Likert scale (1 strongly disagree to 5 strongly agree). To sum the score, for each of the odd-numbered items, we need to subtract one from the score (one, three, five, seven, and nine). For each of the even-numbered questions, we should subtract their value from five (two, four, six, eight, and ten). Then the total score is multiplied in 2.5, which is ranged between 0 and 100. A score of 55% indicates the highest satisfaction and less than 55% indicates dissatisfaction (23). the reliability and validity of the Persian Version of the above scale have been established in a published report (24)

### 2.4. Procedures

The study was done in a quiet room with in the day care centers. Having provided the informed consent, the volunteer participants were individually trained by a researcher how to work with the app. Next, they were asked to work with app without the researcher’s help. Yet, the researcher was available to respond to their possible questions. After completing the test and evaluation of the app, participants underwent neuropsychological testing using the paper-based MoCA battery. The medical ethics committee of Shiraz University of medical science approved the study protocol (Ethical code: IR.SUMS.REC.1397.715).

### 2.5. Statistical Analyses

To evaluate the face validity and SUS, the mean scores were considered. Pearson correlation and Kuder-Richardson were used to evaluate the convergent validity, test-retest reliability, and internal consistency, respectively. Reliability correlation coefficient values higher than 0.7 were considered to be satisfactory. The analysis was done in IBM SPSS software 16.0 and Microsoft Excel. A *p*-value of <0.05 was considered as statistically significant in all statistical procedures.

### 2.6. Security and Privacy concerns

Incorporating health information into the app is an issue which specialists and users are often concerned about since the mobile phone is easily portable and accessible anywhere. To overcome this concern, users of the M-Check app need to first enter a password to reach the interface. Users can also delete the account, or easily change the password.

## 3. Results

### 3.1. Usability evaluation

The average SUS usability evaluation scores were 82.5 and 77.11, obtained from the expert evaluators and the elderly subjects, respectively. According to the usability scale, a score of over 55% is considered as favorable. Therefore, the evaluation score was appropriate as per the report both by the expert and elderly groups.

### 3.2. Sociodemographic characteristics

Mean (Standard Deviation) age of the participants was equal to 65. 9 years old (5.3 years), with an age range of 60-82 years old. Sociodemographic characteristics of study participants are shown in Table 1.

**Table 1.** Sociodemographic characteristics of study participants

| Variables | Patient (n=70)n (%) |
| --- | --- |
| **Sex** |  |
| Male | 49(70) |
| Female | 21(30) |
| **Educational status** |  |
| ≤ 12 | 50(71.4) |
| >12 | 20(28.6) |
| **Left -handed** | 5(7.1) |
| **Right -handed** | 61(87.1) |
| **Both -handed** | 4(5.7) |
| **Smoking** |  |
| Yes | 13(18.6) |
| No | 57(81.4) |
| **Alcohol use** |  |
| Yes | 6(8.6) |
| No | 64(91.4) |
| **Physical activity** |  |
| Yes | 43(61.4) |
| No | 27(38.6) |
| **Family History of Alzheimer disease or forgetfulness** |  |
| Yes | 7(10) |
| No | 63(90) |

### 3.3. Face validity

App’s face validity was completed by experts (n = 6) and elderlies (n = 10) groups. The overall mean face validity for the elderly was 9.52%, and the overall mean face validity of the experts was 8.75.

### 3.4. Convergent Validity

Pearson’s coefficient “r” was used to evaluate the convergent validity. Pearson’s coefficient at this stage was - 0.71. Hence, there was a significant negative correlation between the M-Check app assessment and the total score of MoCA (p <0.05).

### 3.5. Test reliability and test-retest reliability

In the app-based group the internal correlation coefficient was 0.88, indicating the reliability of the tool. On the other hand, the paper-based group indicated the internal correlation coefficient of 0.92, indicating the high reliability of the instrument.

### 3.6. Internal Reliability Testing

The Coder-Richardson coefficient of cognitive impairment was used to assess the intrinsic reliability of the Cognitive Impairment Self-Assessment. Richardson’s Coder coefficient was 0.82 suggesting is an acceptable reliability.

## 4. Discussion

This study described the development of the mobile instrument i.e. M-Check application. The M-Check app was designed to fill the gap in self-assessment of mild cognitive impairment. The tool aimed at to providing an easily conducted and quickly interpretable assessment. It is also meant to provide general information on MCI for the elderlies and their families. Such an application is expected to be of use and benefit for individuals themselves, their family members or caregivers, as well as health care providers.

Studies have shown that mobile health in the elderly can be useful in improving care, personal management and self-efficacy. Such apps are intended to promote individuals’ health-related behaviors such as sleep, diet, physical activity, and mental health, as well as adherence to medication. Mobile health (m-health) can also be useful in disease prevention, lifestyle modification, cardiovascular disease management, and diabetes(18). M-health is therefore a valuable and appropriate tool for the elderly. The use of mobile health technologies in the assessment of cognitive and psychological domains in the elderly has also been shown reliable (19).

Various studies have been conducted in terms of application designs to evaluate cognitive predicaments. These apps were designed based on the conventional testing batteries (25–27). Results from earlier investigations have endorsed the feasibility of using self-administered computerized cognitive tests in older primary care patients, given the increasing reliance on computers by people of all ages (25). As such, the use of computerized or mobile-based apps are expected to be generally feasible for cognitive self-assessment in elderlies.

Older adults are usually less capable than young people in using new technologies such as smartphones, therefore the modules are designed to be comfortable for this group so that they can work with different parts of the app after a brief tutorial.

The SUS scores in both groups of experts and elderlies showed that the M-Check retains a good usability. In that sense, two of the items in the face validity of the app i.e. “the degree of easily use of the app” and “the satisfaction on the M-Check app” acquired the mean score of 9.2 and 9.9, respectively. The expert panel’s score for the above two items were 9.3 and 8.83,respectively, indicating that the app was designed in a way to be easily used by this age group.

The convergent validity evaluation revealed a proper validity between MoCA and the M-Check, which was less than ideal(r= -0.71). In a previous study, the paper-based convergent validity was higher than the application based test. Also, the test-retest correlation coefficient for participants in both groups i.e. paper- and application-based were 0.92 and 0.88, respectively, suggesting a high reliability for the M-Check app.

Aging is a process that affects many functions of the body, including the brain, which leads to cognitive decline (28). Cognitive changes might include declined processing speed, impaired working memory and memory retrieval (29). It is thus expected that aging per se may affect learning new things such as working with the new app often making it a difficult task. This might have possibly affected the correlation of MoCA and M-Check and also paper-based and mobile-based interfaces.

Possin *et al*. showed that the brain health assessment program had good properties and features for identifying mild cognitive impairment. Also, the results showed that all subsets (memory, executive functions and speed, visuospatial skills, and language, and an optional informant survey) had high validity. The optional informant survey subset that was completed by another person increased the sensitivity of identifying neurocognitive disorder. Findings from their study substantiated that diagnostic approaches towards cognitive impairments and dementias could also be implemented in software platforms with acceptable validity and reliability indices (27).

In another stydy, Rents *et al*. showed that a significant relationship between the program designed for the iPad and the results of the C3-PAD test in the clinic (r2 = 0.508, p <0.0001). There was also a moderate correlation between home assessment and standard paper automated tests (r2 = 0.168, p <0.003) (30). According to their results, cognitive tests completed at home could be used as clinical diagnostic measure in the clinic.

Scanlon *et al*. investigated the accuracy of a smartphone-based CCS tool designed to identify cognitive impairment compared to the MoCA test. Based on their report, patients’ CCS scores were significantly lower than healthy individuals (P <0.001), and the CCS score was significantly correlated with the MoCA test (r = 0.78, p <0.01). Their results indicated that CCS can be a means of identifying symptoms of cognitive impairment at a community level (31).

Zorlogo *et al*. designed a mobile test including 33 questions from 14 types of assessment tests in 8 different cognitive functions for mobiles with Android OS. The designed test could show a significant difference between the healthy group and the patient in the areas of functional, visual, memory, focus, orientation. Meanwhile, no significant difference was found in other areas such as language, abstraction and mathematics. Based on their findings, the correlation with the MoCA test was 0.57(26). The correlation coefficient in the present study was 0.71, which is inconsistent with the result of the present study. Nevertheless, the disparity in results could have been due to the type of testing used in the either studies.

In our present research the usability was a key factor besides content validity. While the app usability was evaluated by VAS and SUS checklists, some users even verbally expressed their views: “I would like to work with that again” or “It is a suitable tool for elderlies and their families”. To our surprise one participant added: “I’d like to use the app in the future as regular self-assessment if it gets a bit more complex”.

This study was a pilot study and is worth to be conducted in a large sample size in the future. The study could have also been implemented in two app formats (self-assessment and assessment based on the participant relatives’ opinion), which was the limitation of the present study.

## 5. Conclusion

We validated the M-Check app for the detection of MCI based on a need for cognitive assessment tools that can identify early phases of cognitive decline. The interface takes much shorter time than the typical neuropsychological batteries to deal with. In addition, the app can be administered without a presence of health professionals or trained assistants. Results from the present research showed that this technology-based cognitive assessment tool retains a promising outlook. Meanwhile, additional work need to be underway to ensure that M-Check is ready to launched for use at community level and clinical purposes.

## Data Availability

Authors confirm the availability of all data referred to in the manuscript.

## 6. Ethical Considerations

This study was approved by the Ethics Committee of Shiraz University of Medical Sciences. All participants submitted written informed consents prior to enrollment and were entitled to quit the study process anytime upon their discretion.

## 8. Acknowledgment

This paper is a part of a MSc. thesis of the first author, entitled “Mobile-Based Self-Assessment Tool for Mild Cognitive Impairment (Design and Assessment)”. The authors wish to thank the Welfare Organization of Fars province (Iran) and senior care centers in Shiraz, Iran. We are grateful to all the experts and the elderlies participated in this study.

